# The impact of thermal and auditory unpleasant stimulus on motor imagery in healthy individuals

**DOI:** 10.1101/2025.03.06.25323546

**Authors:** Gabriel Cohen-Aknine, Pionnier Raphaël, Mottet Denis, Arnaud Dupeyron

## Abstract

Motor imagery is the ability to mentally simulate a motor task without actually performing it. Pain is an unpleasant sensory experience that involves different dimensions - sensory-discriminative, motivational-affective, and cognitive-evaluative - that are known to interfere with motor imagery. However, it remains unclear which specific pain dimension most significantly impairs motor imagery.

This study aims to compare the effects of unpleasant auditory (primarily affective and cognitive) and thermal (primarily sensory) stimuli, which can be assimilated to pain, on discrete and continuous explicit motor imagery modalities. Eighteen healthy participants were exposed to unpleasant stimuli in addition to a control condition. Participants rated their motor imagery abilities after tasks involving rest, motor execution, and motor imagery in discrete and continuous wrist movement modalities.

Results showed that during discrete motor imagery, only the aversive auditory stimulus significantly reduced motor imagery abilities, whereas thermal pain had no effect. In contrast, motor imagery abilities were preserved during the continuous modality.

These findings suggest that explicit motor imagery may be more affected by the affective dimension of pain induced by aversive auditory stimuli. The preservation of motor imagery abilities in the continuous modality provides insight into the optimization of rehabilitation programs.

## Introduction

Pain is defined by the IASP (International Association for the Study of Pain) as “an unpleasant sensory and emotional experience associated with actual or potential tissue damage, or described in terms of such damage” (1). It is a multidimensional and dynamic process involving sensory-discriminative (e.g. intensity, localization), motivational-affective (e.g. anxiety, escape strategies) and cognitive-evaluative (e.g. attention, memory) dimensions (2–4).

Exercise is known to reduce pain sensation, a phenomenon referred to as exercise-induced hypoalgesia (5). However, in individuals with chronic pain conditions, the hypoalgesic effects of rehabilitation exercise are often mitigated (6–8). As a result, alternative rehabilitation approaches such as motor imagery (MI) have been proposed. MI has been shown to be beneficial in rehabilitation (9,10), particularly for patients with pain (11,12), as it allows stimulation of the sensorimotor cortex without the pain associated with actual movement.

Motor imagery is the ability to imagine performing a motor task without actually doing it (13). It can take several forms, including implicit MI, which involves mentally rotating and recognizing images of the body (14,15), and explicit MI, which involves imagining the movement of a body part without actually performing the movement. Motor imagery activates regions of the brain that use similar substrates to those involved in sensorimotor or motor (20–22).

However, some studies suggest that the experience of pain may reduce MI abilities in individuals with chronic pain (23,24), although these findings vary depending on the type of pain (25). In experimental pain research, pain can be induced by various stimuli, such as thermal pain (26,27). Based on the IASP definition of pain as an unpleasant experience, pain-like experiences have been observed with aversive auditory stimuli that produce pain intensities comparable to thermal pain and activate common brain patterns associated with negative valence and affective states (28–30). A previous study showed that experimental pain induced by electrical stimulation could interfere with MI processes in healthy individuals (31). However, to our knowledge, little is known about whether the type of pain and its different dimensions (affective, cognitive and sensory) influence MI abilities.

In addition, the characteristics of the movement required to the patient can be disruptive. Movements can generally be performed in discrete (a single full or limited range of motion) or continuous (repeated full or limited range of motion) modalities (32). Sensorimotor brain patterns are differentially activated by discrete and continuous movement modalities (33,34). Similarly, sensorimotor activity has been shown to vary between discrete and continuous MI modalities (35). However, the effects of experimental unpleasant stimuli on discrete versus continuous MI modalities remain poorly understood.

The aim of this study was to compare the effects of unpleasant thermal and auditory stimuli with a control resting state condition on explicit motor imagery abilities in healthy subjects during discrete and continuous modalities. Based on previous findings, we hypothesized that MI abilities would be reduced under unpleasant conditions regardless of the MI modality (discrete or continuous).

## Material and methods

This work is part of a larger, experimental single center prospective design study conducted at the Research Unit of Euromov Digital Health & Motion (Montpellier, France). The study was approved by the local ethics committee (Comité d’Éthique de la Recherche de l’Université de Montpellier: n° UM 2023-031) according to the declaration of Helsinski revised in 2013. Participants received and signed a written informed consent form.

### Participants

Eighteen healthy right-handed participants, aged 18-50 years, with no history of neurological or psychiatric disorders, no chronic pain and no pain in the upper limbs during the experiment were included. Students and staff of the University of Montpellier were recruited to participate. Recruitment was carried out by email and posters around the university. Participants were excluded from the study if they were unable to perform the procedure described below and/or because of a dysfunction of any equipment used in the protocol. The inclusion period has been realized between November 20, 2023 and February 22, 2024.

Age, sex, weight, height, and body mass index (BMI) were recorded. Participants’ handedness was assessed using the Edinburgh Handedness Inventory - Short Form (EHI-SF) (36), a questionnaire that scores handedness based on 10 daily activities, with a ratio and a cut-off of 40 points or more indicating right-handedness. Levels of physical activity were measured using the International Physical Activity Questionnaire-Short Form (IPAQ-SF) (37). This self-report questionnaire uses an algorithm to calculate the number of minutes of physical activity (PA) per week, categorized into three levels (low, moderate and vigorous), and a score for inactivity (corresponding to the number of minutes spent sitting). The total score is expressed in MET (Metabolic Equivalent of Task) minutes/week defined as the ratio of the energy expended during the activity in question to the amount of energy expended at rest. For example, 1 MET corresponds to resting metabolic rate or basal metabolic rate (38).

Participants’ explicit motor imagery ability was measured using the Kinesthetic and Visual Imagery Questionnaire (KVIQ) (39). This questionnaire consists of 14 items: 7 for visual motor imagery (VMI), i.e., imagining the visualization of movements and 7 for kinesthetic motor imagery (KMI), i.e., imagining the sensation of movements. Participants were asked to perform different movements, then imagine them and rate their imagery abilities on a 5-point Likert scale (1 = no image/no sensation to 5 = image as clear as seeing/as intense as performing the action).

### Procedure

Participants were seated in a chair in front of a computer in a relaxed position with their forearms resting on the armrests and their elbows naturally flexed in pronation, so that the wrists were fully flexed and relaxed. The procedure consisted of 2 parts: a discrete modality (consisting of single imagined and executed wrist movements) and a continuous modality (consisting of repeated imagined and executed wrist movements). The instructions were displayed on a screen using PsychoPy software (v 2023.1.2) (40,41).

In the discrete modality, three tasks were performed: “resting state”, “motor execution” and “motor imagery”, and only “motor execution” and “motor imagery” for the continuous assessment. During the motor execution and imagery tasks, a blank screen was displayed between the on-screen ‘GO’ and ‘STOP’ instructions. During the resting state, a central cross was displayed between the ‘GO’ and ‘STOP’ instructions on the screen. Before each task, general instructions were displayed to remind participants of the requirements of each task. For example, before the resting state task, the instructions to fix the cross and stay relaxed were displayed.

For the “motor execution” task, participants were instructed to perform the movement described below, in parts 1 and 2. For the motor imagery task, they were asked to imagine performing the same movement as in the “motor execution” task, with no specific instructions regarding visual or kinesthetic motor imagery modalities. To familiarize participants with the movement, a training session was conducted prior to the experiment in which participants were asked to perform the movement “in their own way” with respect to the previous instructions, without additional guidance.

#### Part 1: discrete

With the wrist fully flexed as described above, participants were instructed to perform a full range of motion wrist movement in the sagittal plane (from fully flexed to fully extended position), known as discrete modality, and return to the starting position, within a time window of 4 seconds. This procedure was repeated 20 times.

#### Part 2: continuous

In the same position as part 1, participants were instructed to perform a repetitive/continuous full range of motion wrist movement in the sagittal plane (from fully flexed to fully extended position) for 25 seconds. This procedure was repeated 3 times.

The two painful conditions and control condition were tested for both parts.

### Conditions

Three blocks of conditions were created: ‘normal’ (= control), ‘auditory’ (aversive auditory stimulus) and ‘heat’ (thermal painful stimulus). The order of the conditions was pseudo-randomised using six task sequence designs (e.g., normal, auditory, heat; heat, auditory, normal…) determined by dice. In the ‘normal’ condition, movement was performed and imagined without any unpleasant stimulus.

The aversive ‘auditory’ stimulus was created using Audacity® (v 3.3.3) with a duration of 4 seconds, a start and end fade of 0.2 seconds, a frequency of 5000 Hz (sawtooth waveform) and a baseline volume of 75 dB. This stimulus was inspired by a previous publication by Valentini et al (29). The thermal ‘heat’ stimulus was applied using hot water in a 14/19 L thermostatic bath (CORIO C-BT19, Julabo®, Seelbach, Germany) with a baseline temperature of 45°C (42).

To standardize variability between participants, each participant selected their level of discomfort for each stimulus on a 100-point visual analogue scale (VAS) (0 = ‘no discomfort’ to 100 = ‘unbearable discomfort’). Participants were instructed to choose the water temperature and the auditory sound within a range between 60 and 75 on the VAS (Fig. **1**). The levels were adjusted in increments of 1°C for temperature and 1dB for sound volume until the chosen level of discomfort was reached.

**Fig. 1.**
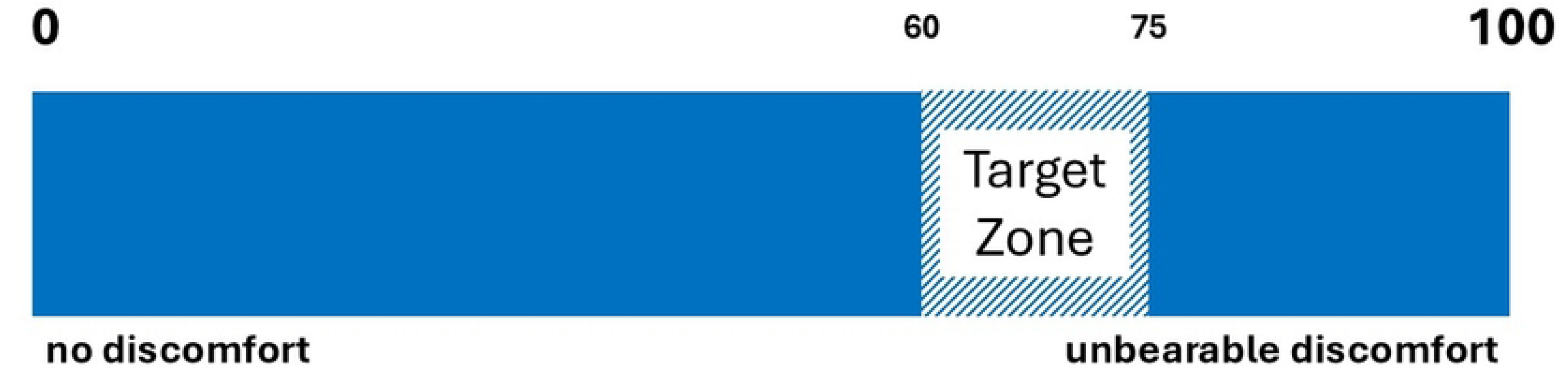
Analogue visual scale for unpleasant stimulus levels.

For the thermal stimulus, the right hand was immersed in water up to the wrist joint, and for the auditory stimulus, the sound was played for periods of 25 seconds through loudspeakers. The temperature level was monitored using the CORIO C-BT19 thermometer and the aversive auditory level was measured using the “Decibel X - Pro Sonomètre smartphone application” (SkyPaw Co.®, Ltd) (43). A thermal heat stimulus was chosen because it results in a shorter duration of pain, a more constant quantitative description and a steeper slope for the intensity of the sensation compared to cold pain stimulus (44). The design of the protocol and the instructions are shown in Figs 2 and 3 respectively.

**Fig. 2.**
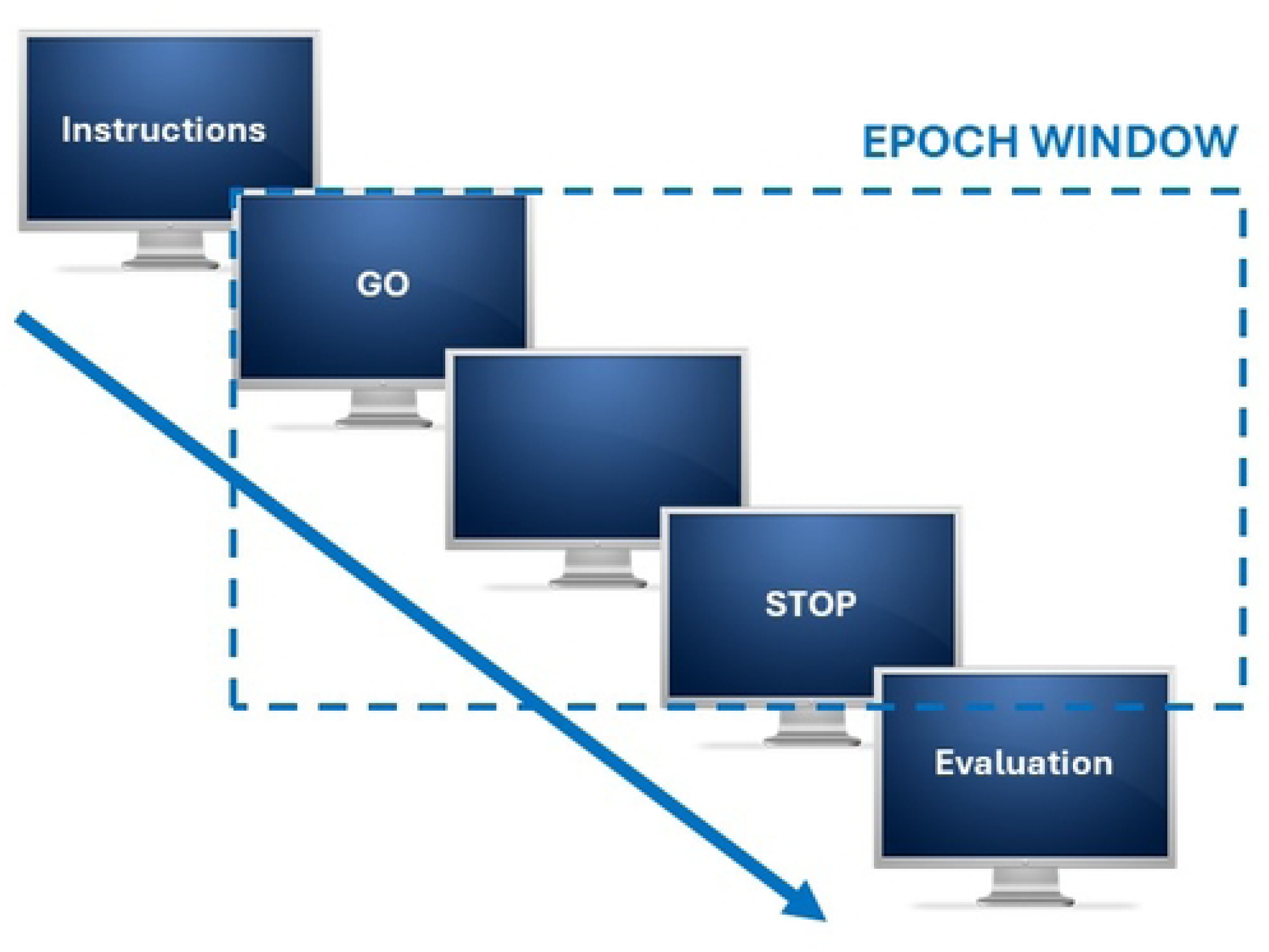
Instructions design.

**Fig. 3.**
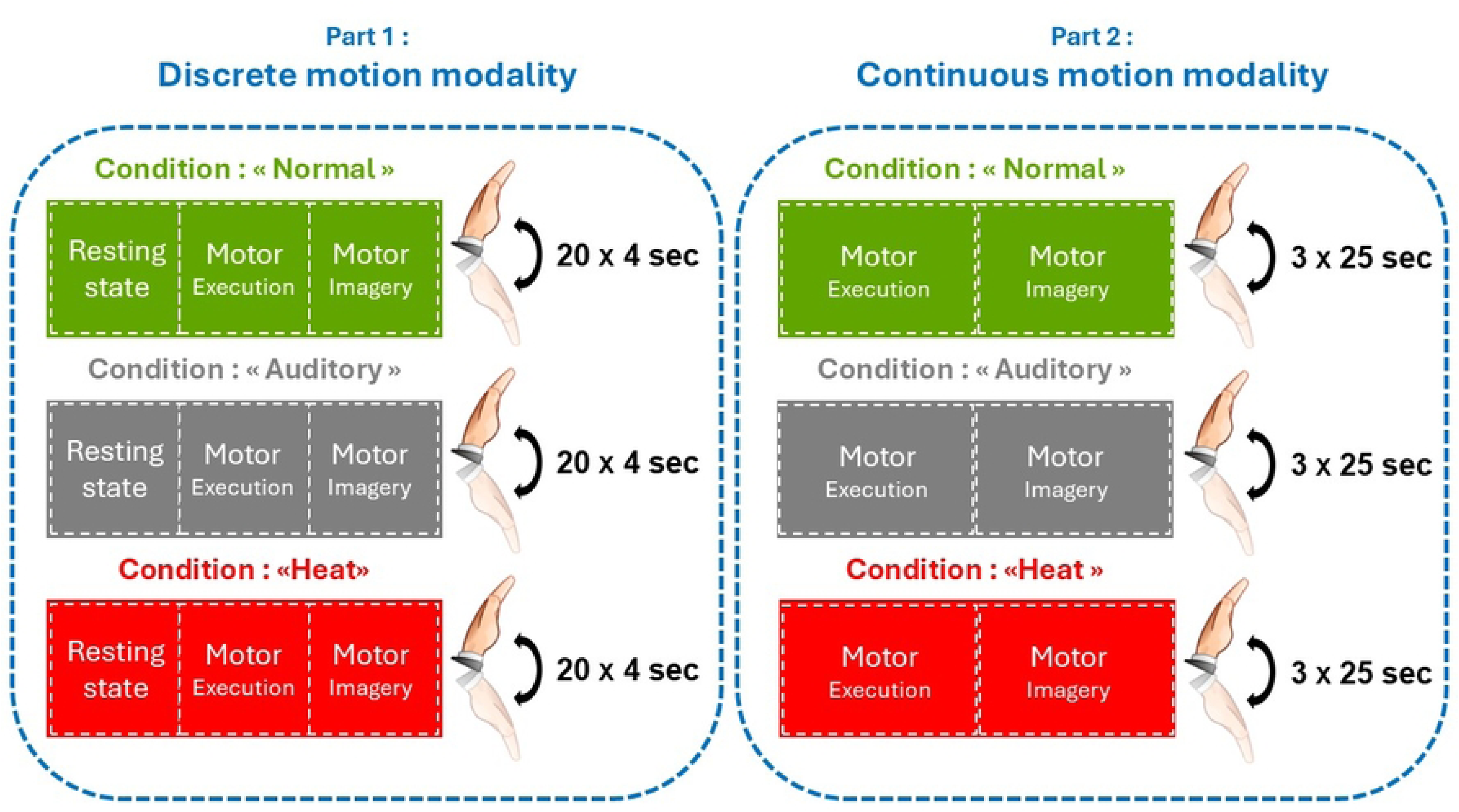
Protocol design.

**Fig. 4.**
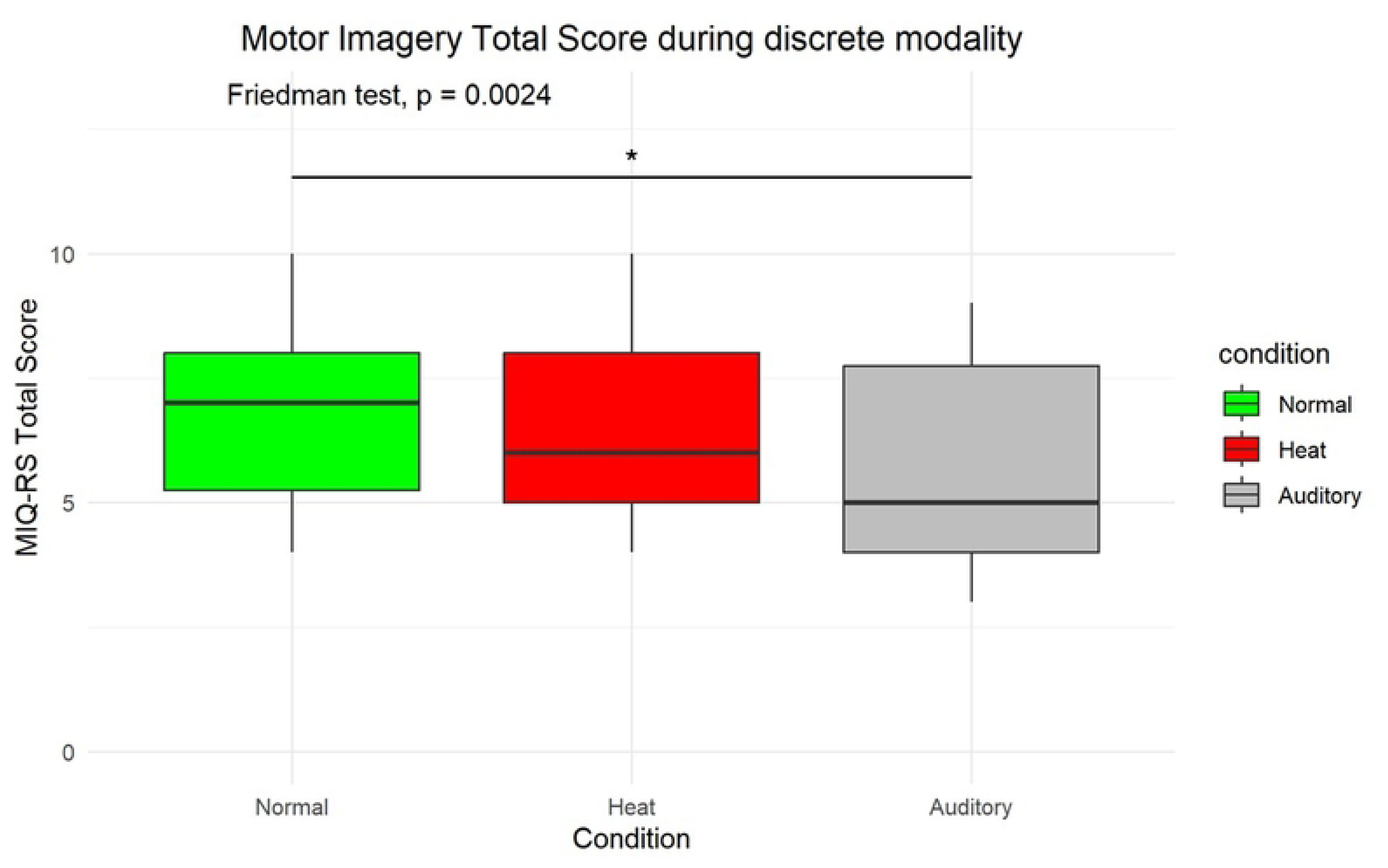
Box plot of motor imagery scores between Normal, Aversive Auditory and Thermal Heat conditions during the discrete motor imagery modality.

**Fig. 5.**
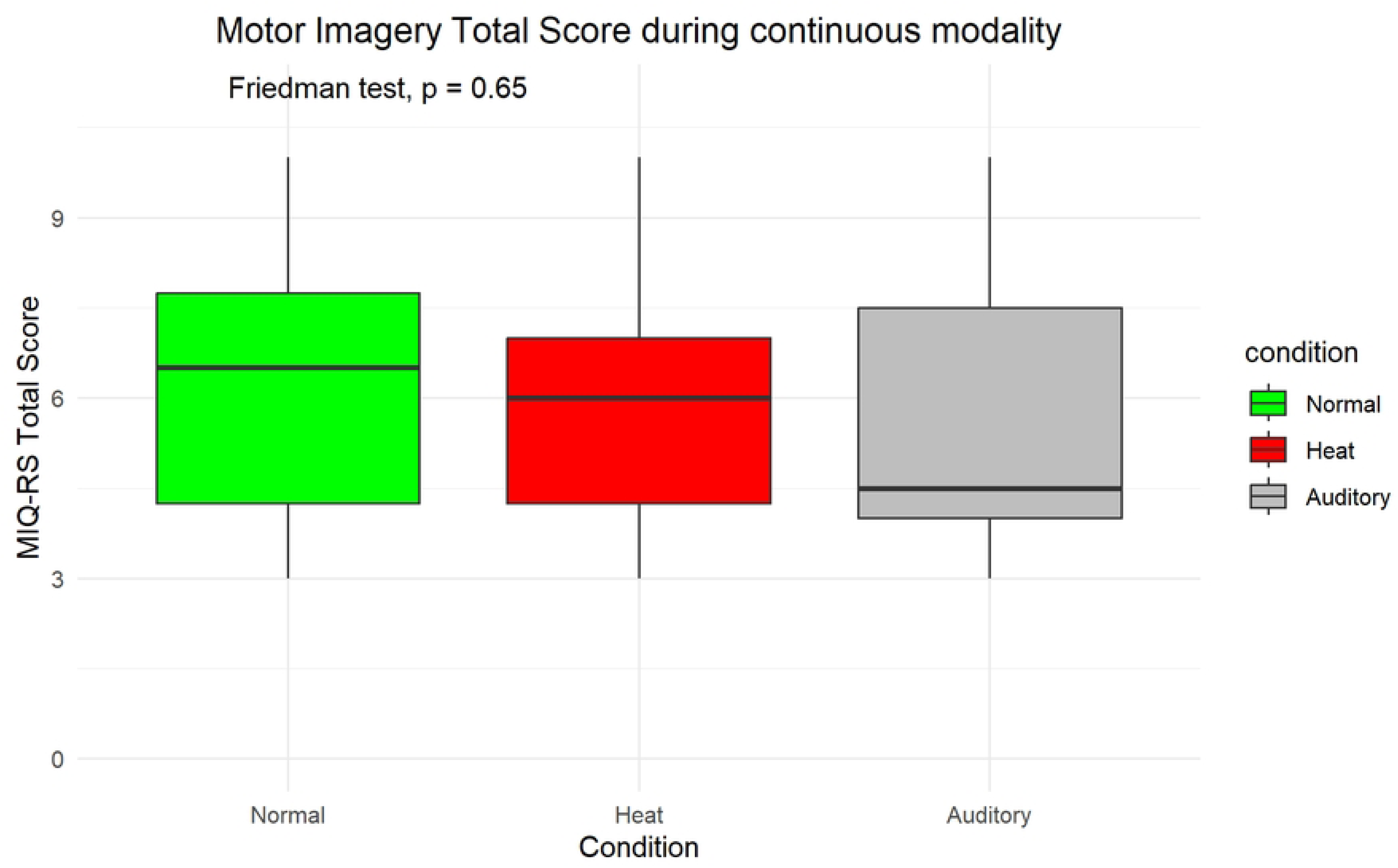
Box plot of motor imagery scores between Normal, Aversive Auditory and Thermal Heat conditions during the continuous motor imagery modality.

### Primary outcome measures

The effect of pain sensation on motor imagery ability was assessed using the 5-point Likert scale of the KVIQ. The two kinesthetic (KMI) and visual (VMI) motor imagery subscales were recorded at the end of each task condition (normal, heat and auditory) for each modality (discrete and continuous), and the total score was composed of both subscales.

### Statistical analysis

Statistical analyses were performed using R Studio® software (v. 2024.04.2). Due to the small sample (45,46), the Friedman test was used to compare conditions, and post hoc analysis was performed using the paired Wilcoxon test with the Bonferroni-Holm correction to adjust for multiple comparisons. Outcome data were reported as median (interquartile range) and confidence intervals with upper and lower limits.

Effect sizes for statistically significant differences between conditions were calculated using Kendall’s tau. Statistics are presented with a 95% confidence interval and an alpha risk of 0.05.

## Results

### Demographic analysis

Eighteen participants (7 females and 11 males) were included in the study. The descriptive analysis is detailed in the table 1.

**Table 1.** Descriptive characteristics of the participants.

| Variable | Median | IQR | CI_upper | CI_lower |
| --- | --- | --- | --- | --- |
| Age (years) | 18.50 | 1.75 | 20.83 | 16.91 |
| Size (meter) | 177.50 | 7.00 | 184.09 | 168.41 |
| Mass (kg) | 75.50 | 18.50 | 92.97 | 51.53 |
| BMI | 23.35 | 3.80 | 27 | 18.49 |
| IPAQ score | 4 290 | 4 305.75 | 10 416.98 | 773.77 |
| Inactivity score | 2 730 | 1 522.50 | 4 251.15 | 841.34 |
| KVIQ VMI | 18 | 2 | 21.24 | 16.76 |
| KVIQ KMI | 18.50 | 1.75 | 20.83 | 16.91 |
| Volume (dB) | 81 | 7.75 | 87.55 | 70.20 |
| Temperature (°C) | 46 | 1.88 | 48.04 | 43.84 |

CI: Confidence interval

### Primary outcome: Motor Imagery ability

All details of motor imagery statistical analysis are provided in Appendix 1.html.

#### Discrete motor imagery modality

There were significant statistical differences between conditions (p = 0.002). Post-hoc analysis revealed that the auditory stimulus decreased motor imagery abilities compared to the normal condition (p = 0.001) with a large effect size (d = 0.703) (Table 3. **Summary of the Total MIQ-RS score in each condition during the continuous motor imagery modality**). Summaries are presented in Table 2.

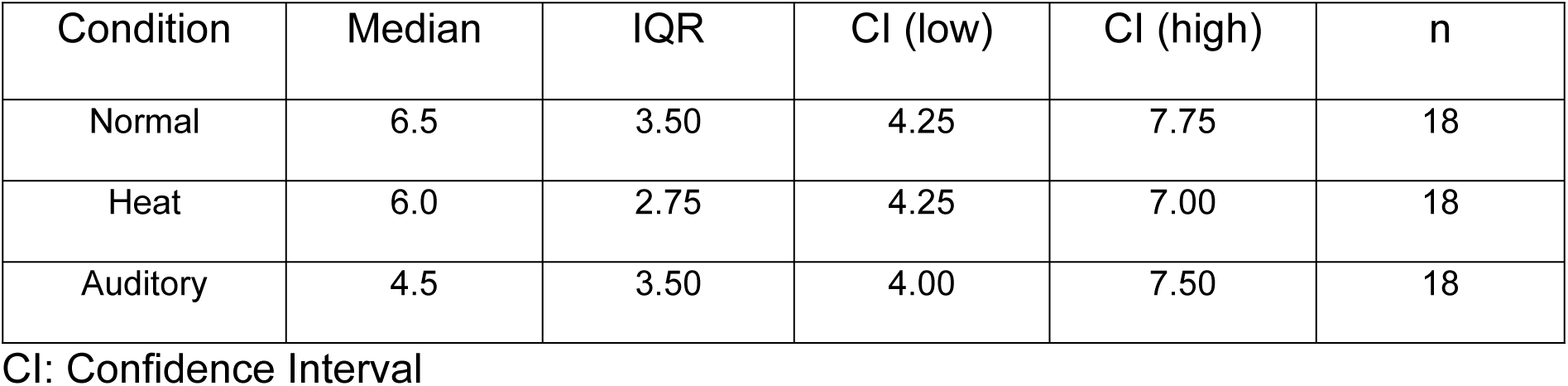

**Table 2.** Summary of the Total MIQ-RS score in each condition during the discrete motor imagery modality.

| Condition | Median | IQR | CI (low) | CI (high) | n |
| --- | --- | --- | --- | --- | --- |
| Normal | 7 | 2.75 | 4.323 | 9.677 | 18 |
| Heat | 6 | 3.00 | 4 | 9.354 | 18 |
| Auditory | 5 | 3.75 | 3 | 8.677 | 18 |

CI: Confidence Interval

**Table 3.**
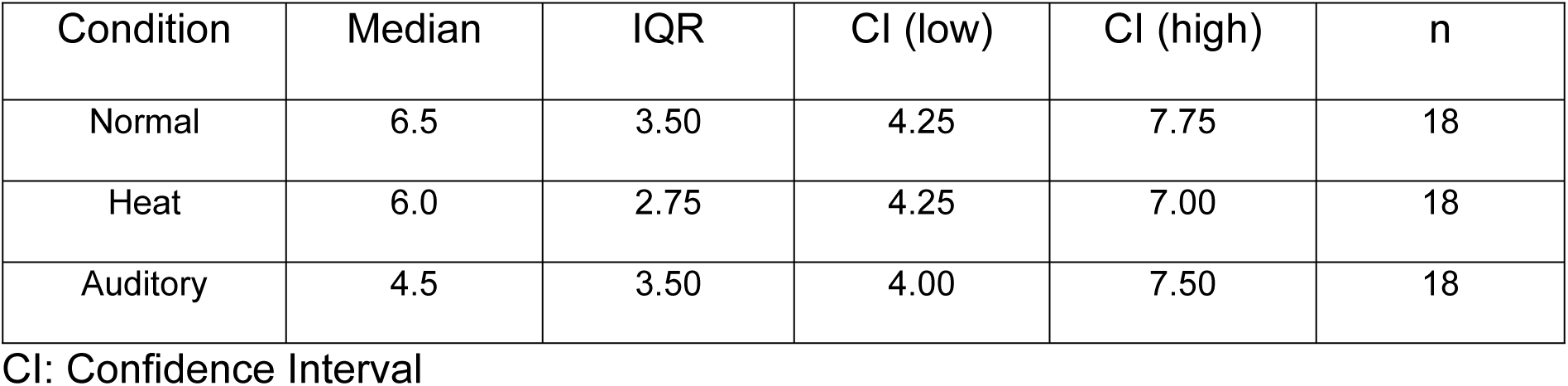
Summary of the Total MIQ-RS score in each condition during the continuous motor imagery modality.

#### Continuous motor imagery modality

There were no significant statistical differences between conditions (p = 0.645), indicating that the unpleasant conditions did not impair motor imagery during the continuous motor imagery task (**Error! Reference source not found.**). Summaries are presented in Table 3.

## Discussion

Our hypothesis that motor imagery abilities would be reduced by unpleasant stimuli was partially confirmed, as only the aversive auditory stimulus decreased motor imagery scores, specifically during the discrete motor imagery modality. However, for the continuous motor imagery task, our hypothesis was not confirmed for either unpleasant stimulus.

Firstly, consistent with previous study (47), we found a decrease in motor imagery abilities during the aversive auditory stimulus, but only during the discrete modality, not the continuous one. Some authors have postulated that motor imagery may be more influenced by affective dimensions of pain, such as cognitive-evaluative or motivational-emotional factors (48). In our study, we used two unpleasant stimuli to differentiate distinct aspects of pain, contrasting an affective stimulus without a localized component (aversive auditory stimulus) with a conventional experimental pain model (thermal heat stimulus). Our results suggest that motor imagery abilities may be more influenced by the affective dimension of pain than to its sensory-discriminative dimension. Motor imagery is part of the Movement Representation Methods (MRM) (12), and previous research has shown that explicit motor imagery is more cognitively demanding than other techniques within this therapeutic approach (48). In addition, studies have shown that motor imagery relies on working memory storage and induces mental fatigue (49) whether during prolonged implicit (50) or explicit motor imagery (51). Furthermore, research has shown a relationship between working memory and the perception of aversive sensations in response to unpleasant stimuli (52,53). Some researchers have compared aversive auditory stimuli to a tinnitus-like sensation, which has been shown to activate cortical areas involved in attention and negative emotional responses (54). Our results suggest that the affective dimension of pain may impair explicit motor imagery. Therefore, given the literature indicating that cognitive demands are high during explicit motor imagery tasks, and given the relationships between aversive sensations and working memory, it is plausible that the reduction in motor imagery abilities observed in our study was induced by the aversive auditory stimulus due to its ability to disrupt attentional processes such as working memory.

These disturbances were observed only in the discrete motor imagery modality, not in the continuous modality. It has been suggested that brain activity in cortical motor planning areas increases during discrete versus continuous motor tasks (33), with greater attentional demands in the discrete modality (32). In support of this, previous research has shown that continuous movements are less affected by cognitive load than discrete movements (34). In our study, the discrete condition consisted of 60 trials (3 × 20 tasks) of discrete explicit motor imagery, whereas the continuous condition consisted of 9 trials (3 × 3 tasks) of continuous explicit motor imagery. The high cognitive demands associated with explicit motor imagery, particularly in the discrete modality, combined with the impact of the aversive auditory stimulus on the attentional network, may explain our findings. This may make the discrete modality more sensitive to the affective-motivational and cognitive-evaluative dimensions of pain.

Regarding the lack of effects of the thermal heat pain stimulus on motor imagery, our results contrast with previous studies suggesting that experimental pain impairs the motor imagery process (47). However, different experimental pain models have been used in the literature, including phasic pain (short duration with intense sensation) and tonic pain (long duration with moderate sensation). For example, a previous study reported a decrease in corticospinal excitability during electrical pain stimulation while participants performed explicit motor imagery of digit V (47), whereas our experimental pain model used a thermal heat pain stimulus. Although thermal and electrical stimuli target nociceptors, research suggests that different experimental pain models engage multiple dimensions of the participant’s experience, with no significant correlation observed between responses to electrical and thermal pain stimuli (26). This suggests that experimental pain models may activate different sensory and affective pathways, resulting in unique individual experiences.

Some reviews have highlighted the impact of pain conditions on implicit motor imagery performance, showing a decrease in accuracy and response times (23,55), regardless of pain localization. Accordingly, some authors have suggested that changes in motor imagery are specifically related to a painful body part (56) likely due to changes in internal body representation (57). However, other studies have shown that patients with widespread pain, such as fibromyalgia, have reduced implicit motor imagery abilities (58,59), suggesting a more complex interaction between pain localization and motor imagery deficits. Additionally, a study of women with menstrual pain demonstrated a reduction in explicit motor imagery abilities, assessed with the same questionnaire as in our study, with results showing a negative correlation between non-localized unpleasant sensory, such as somatic complaints related to menstrual symptoms, and motor imagery scores (60). In our study, we used an aversive auditory stimulus, a non-localized unpleasant sensory input, and found that explicit motor imagery performance was more significantly impaired than that observed with a localized thermal unpleasant stimulus. These results suggest that the localization of the unpleasant stimulus alone does not fully explain motor imagery deficits. Indeed, another study highlight the role of the affective dimension in motor imagery performance, reinforcing the idea that motivational-affective, and cognitive-evaluative factors interact to influence motor imagery process (61).

A complementary analysis supported our interpretation. First, the motor imagery abilities can be subdivided into two dimensions as kinesthetic (KMI is related to the sensation of imagined movement) and visual (VMI is related to the visualization of the imagined movement) motor imagery subscores. Our complementary analysis supported our results for the KMI and VMI for the discrete and continuous motor imagery modalities. Second, the intensity of the unpleasant sensation after induced stimulus was equivalent in the aversive auditory and heat pain stimulus compared to the control condition whether during discrete and continuous motor imagery modalities. These results are consistent with previous studies (29,62). Finally, a correlation analysis was performed and showed a negative moderate to strong correlation between unpleasantness intensity and explicit motor imagery abilities for the aversive auditory stimulus but not the thermal heat pain stimulus, confirming our findings regarding the nondisruptive effect of localized stimuli on explicit motor imagery abilities (complete analysis is provided in the Appendix 1.html). This suggests that the affective and cognitive dimension of pain is more likely to decrease explicit motor imagery process.

### Clinical implications

Our results showed that the continuous motor imagery modality was not impaired by unpleasant stimuli. These findings support the use of motor imagery as a valuable therapeutic tool, as self-reported motor imagery abilities appear to be preserved in the presence of pain. This highlights an opportunity to optimize the implementation of motor imagery exercises and reinforces its potential as a rehabilitation strategy, given its demonstrated efficacy in reducing pain and improving range of motion (63). Therefore, explicit motor imagery therapy should be prioritized within a continuous exercise modality, while considering the risk of mental fatigue associated with the cognitive demands of motor imagery tasks, as discussed previously. However, our results were assessed in an observational design study, longitudinal evaluation should be tested to give a more robust therapeutic dimension of our results. In addition, our results suggest that the affective dimension of pain, assessed with an aversive auditory stimulus, impairs motor imagery abilities specifically in the discrete modality. Previous studies have shown that psychosocial aspects of pain influence quality of life in interdisciplinary pain rehabilitation (64). Thus, assessment of the affective dimension of pain (65) may be important prior to initiating rehabilitation programs, particularly those involving motor imagery in a discrete modality.

### Limitations

First, our results were based on a small sample size, which may have limited our ability to capture the influence of gender, a factor known to influence the temporal aspects of explicit motor imagery (66). However, a previous factor analysis of several experimental pain responses indicated that gender did not influence thermal heat pain and that 20% of the variance in pressure pain threshold was explained by heat pain sensitivity, supporting the reliability of our findings (26). In addition, our sample consisted of relatively young participants with a median age of 18.5 years. As age has been shown to influence motor imagery abilities (66–68), this may have introduced a potential bias in our results.

Another potential limitation of our study is that the participants had high levels of physical activity (15 of 18 participants exceeded 3000 MET/min/week, classified as vigorous physical activity), which may not be representative of the general population. A meta-analysis has shown that athletes have better tolerance to experimental pain (69) and that individuals with high motor expertise, such as athletes, rely on different neural mechanisms during motor imagery and are considered as good imagers (70).

Third, the literature includes a wide variety of motor imagery assessment tools, ranging from clinical measures such as the Lateral Judgment Task for implicit motor imagery (71), and mental chronometry (72) to self-report questionnaires for explicit motor imagery (73), as well as neuroimaging techniques such as electroencephalography (74) and functional magnetic resonance imaging (75). This methodological diversity may have limited our conclusions and requires caution in generalizing our findings.

Finally, expectations of positive or negative experiences, commonly referred to as placebo or nocebo effects, suggest that an individual’s predictive cognition of unpleasant stimuli could introduce bias into our study (76). However, previous research has shown that volunteers participating in experimental pain studies tend to be less sensitive to pain and exhibit less fear of pain (77). This may mitigate potential bias, as any observed differences in individuals with lower pain sensitivity suggest that the effect size may be even larger in the general population.

### Opportunities

Recent research on corticospinal excitability suggests that the effects of experimental pain differ from those of clinical pain, highlighting the need for new experimental pain models to better understand how clinical pain affects motor bevahior (78). In particular, aversive auditory stimuli may provide an opportunity to disrupt attentional processes in ways similar to those observed in patients with chronic pain (79). Furthermore, while experimental pain models have been shown to replicate the spatial distribution of pain, they often fail to capture the qualitative aspects of pain as assessed by tools such as the McGill Pain Questionnaire (80). Therefore, combining aversive auditory stimuli with thermal heat pain may provide a more comprehensive model that incorporates additional affective and cognitive dimensions of pain (26).

## Conclusion

This study shows that motor imagery abilities in healthy individuals are differentially affected by the type of unpleasant stimulus and task modality. The motivational-affective and cognitive-evaluative dimensions of pain are more likely to impair discrete motor imagery compared to thermal pain, despite similar levels of unpleasantness. However, when performed in a continuous modality, motor imagery abilities are preserved. These findings suggest that continuous motor imagery may be a more effective therapeutic approach, although attention must be paid to fatigue and working memory disturbances. Future research could explore the combination of aversive auditory and conventional pain models in a longitudinal design.

## Data Availability

The data underlying the results presented in the study are available from an excel.

## Acknowledgments

We would like to thank the 2 research engineers Pierre Jean and Simon Plat from Euromov DHM for their help with software installation, logistics and signal analysis.

## BIBLIOGRAPHY

1. Raja SN, Carr DB, Cohen M, Finnerup NB, Flor H, Gibson S, et al. The Revised IASP definition of pain: concepts, challenges, and compromises. Pain. 1 sept 2020;161(9):1976-82.

2. Kucyi A, Davis KD. The Neural Code for Pain: From Single-Cell Electrophysiology to the Dynamic Pain Connectome. Neuroscientist. août 2017;23(4):397-414.

3. Cholewicki J, Popovich JM, Aminpour P, Gray SA, Lee AS, Hodges PW. Development of a collaborative model of low back pain: report from the 2017 NASS consensus meeting. Spine J. juin 2019;19(6):1029-40.

4. Murray GM, Sessle BJ. Pain-sensorimotor interactions: New perspectives and a new model. Neurobiology of Pain. 1 janv 2024;15:100150.

5. Tanaka K, Kuzumaki N, Hamada Y, Suda Y, Mori T, Nagumo Y, et al. Elucidation of the mechanisms of exercise-induced hypoalgesia and pain prolongation due to physical stress and the restriction of movement. Neurobiol Pain. 2023;14:100133.

6. Polaski AM, Phelps AL, Kostek MC, Szucs KA, Kolber BJ. Exercise-induced hypoalgesia: A meta-analysis of exercise dosing for the treatment of chronic pain. PLOS ONE. 9 janv 2019;14(1):e0210418.

7. Rice D, Nijs J, Kosek E, Wideman T, Hasenbring MI, Koltyn K, et al. Exercise-Induced Hypoalgesia in Pain-Free and Chronic Pain Populations: State of the Art and Future Directions. The Journal of Pain. 1 nov 2019;20(11):1249-66.

8. Wewege MA, Jones MD. Exercise-Induced Hypoalgesia in Healthy Individuals and People With Chronic Musculoskeletal Pain: A Systematic Review and Meta-Analysis. J Pain. janv 2021;22(1):21-31.

9. Harris J, Hebert A. Utilization of motor imagery in upper limb rehabilitation: a systematic scoping review. Clin Rehabil. nov 2015;29(11):1092-107.

10. Ladda AM, Lebon F, Lotze M. Using motor imagery practice for improving motor performance – A review. Brain and Cognition. 1 juin 2021;150:105705.

11. Cuenca-Martínez F, Reina-Varona Á, Castillo-García J, La Touche R, Angulo-Díaz-Parreño S, Suso-Martí L. Pain relief by movement representation strategies: An umbrella and mapping review with meta-meta-analysis of motor imagery, action observation and mirror therapy. Eur J Pain. févr 2022;26(2):284-309.

12. Thieme H, Morkisch N, Rietz C, Dohle C, Borgetto B. The Efficacy of Movement Representation Techniques for Treatment of Limb Pain—A Systematic Review and Meta-Analysis. The Journal of Pain. févr 2016;17(2):167-80.

13. Moran A, Guillot A, MacIntyre T, Collet C. Re-imagining motor imagery: Building bridges between cognitive neuroscience and sport psychology. British Journal of Psychology. 2012;103(2):224-47.

14. Brusa F, Erden MS, Sedda A. More implicit and more explicit motor imagery tasks for exploring the mental representation of hands and feet in action. Exp Brain Res. 1 déc 2023;241(11):2765-78.

15. Conson M, De Bellis F, Baiano C, Zappullo I, Raimo G, Finelli C, et al. Sex differences in implicit motor imagery: Evidence from the hand laterality task. Acta Psychol (Amst). févr 2020;203:103010.

16. Lee WH, Kim E, Seo HG, Oh BM, Nam HS, Kim YJ, et al. Target-oriented motor imagery for grasping action: different characteristics of brain activation between kinesthetic and visual imagery. Sci Rep. 4 sept 2019;9(1):12770.

17. Monsma EV, Gregg MJ, Seiler B, Sacko RS, Hall CR. Convergent validity and sex invariant factor structure of the Movement Imagery Questionnaire-3 - Second version (MIQ-3S): Healthy, young adult reference data. Musculoskelet Sci Pract. juin 2022;59:102537.

18. McInnes K, Friesen C, Boe S. Specific Brain Lesions Impair Explicit Motor Imagery Ability: A Systematic Review of the Evidence. Archives of Physical Medicine and Rehabilitation. mars 2016;97(3):478–489.e1.

19. Vasilyev A, Liburkina S, Yakovlev L, Perepelkina O, Kaplan A. Assessing motor imagery in brain-computer interface training: Psychological and neurophysiological correlates. Neuropsychologia. mars 2017;97:56-65.

20. Hardwick RM, Caspers S, Eickhoff SB, Swinnen SP. Neural correlates of action: Comparing meta-analyses of imagery, observation, and execution. Neurosci Biobehav Rev. nov 2018;94:31-44.

21. Confalonieri L, Pagnoni G, Barsalou LW, Rajendra J, Eickhoff SB, Butler AJ. Brain Activation in Primary Motor and Somatosensory Cortices during Motor Imagery Correlates with Motor Imagery Ability in Stroke Patients. ISRN Neurol. 29 déc 2012;2012:613595.

22. Decety J. Do imagined and executed actions share the same neural substrate? Cognitive Brain Research. 1 mars 1996;3(2):87-93.

23. Breckenridge JD, Ginn KA, Wallwork SB, McAuley JH. Do People With Chronic Musculoskeletal Pain Have Impaired Motor Imagery? A Meta-analytical Systematic Review of the Left/Right Judgment Task. J Pain. févr 2019;20(2):119-32.

24. La Touche R, Grande-Alonso M, Cuenca-Martínez F, Gónzález-Ferrero L, Suso-Martí L, Paris-Alemany A. Diminished Kinesthetic and Visual Motor Imagery Ability in Adults With Chronic Low Back Pain. PM&R. 2019;11(3):227-35.

25. Wallwork SB, Leake HB, Peek AL, Moseley GL, Stanton TR. Implicit motor imagery performance is impaired in people with chronic, but not acute, neck pain. PeerJ. 14 févr 2020;8:e8553.

26. Neziri AY, Curatolo M, Nüesch E, Scaramozzino P, Andersen OK, Arendt-Nielsen L, et al. Factor analysis of responses to thermal, electrical, and mechanical painful stimuli supports the importance of multi-modal pain assessment. PAIN. mai 2011;152(5):1146.

27. Reddy KS kumar, Naidu MUR, Rani PU, Rao TRK. Human experimental pain models: A review of standardized methods in drug development. J Res Med Sci. juin 2012;17(6):587-95.

28. Royet JP, Zald D, Versace R, Costes N, Lavenne F, Koenig O, et al. Emotional Responses to Pleasant and Unpleasant Olfactory, Visual, and Auditory Stimuli: a Positron Emission Tomography Study. J Neurosci. 15 oct 2000;20(20):7752-9.

29. Valentini E, Halder S, McInnerney D, Cooke J, Gyimes IL, Romei V. Assessing the specificity of the relationship between brain alpha oscillations and tonic pain. NeuroImage. 15 juill 2022;255:119143.

30. Čeko M, Kragel PA, Woo CW, López-Solà M, Wager TD. Common and stimulus-type-specific brain representations of negative affect. Nat Neurosci. juin 2022;25(6):760-70.

31. Neige C, Lebon F, Mercier C, Gaveau J, Papaxanthis C, Ruffino C. Pain, No Gain: Acute Pain Interrupts Motor Imagery Processes and Affects Mental Training-Induced Plasticity. Cerebral Cortex (New York, NY: 1991). janv 2022;32(3):640-51.

32. Habas C, Cabanis EA. Neural correlates of simple unimanual discrete and continuous movements: a functional imaging study at 3 T. Neuroradiology. 1 avr 2008;50(4):367-75.

33. Hatsopoulos N, Joshi J, O’Leary JG. Decoding Continuous and Discrete Motor Behaviors Using Motor and Premotor Cortical Ensembles. Journal of Neurophysiology. août 2004;92(2):1165-74.

34. Maes PJ, Wanderley MM, Palmer C. The role of working memory in the temporal control of discrete and continuous movements. Exp Brain Res. 1 janv 2015;233(1):263-73.

35. Rimbert S, Lindig-León C, Fedotenkova M, Bougrain L. Modulation of beta power in EEG during discrete and continuous motor imageries. In: 2017 8th International IEEE/EMBS Conference on Neural Engineering (NER) [Internet]. 2017 [cité 7 oct 2024]. p. 333-6. Disponible sur: https://ieeexplore.ieee.org/document/8008358

36. Veale JF. Edinburgh Handedness Inventory - Short Form: a revised version based on confirmatory factor analysis. Laterality. 2014;19(2):164-77.

37. Meh K, Jurak G, Sorić M, Rocha P, Sember V. Validity and Reliability of IPAQ-SF and GPAQ for Assessing Sedentary Behaviour in Adults in the European Union: A Systematic Review and Meta-Analysis. Int J Environ Res Public Health. 26 avr 2021;18(9):4602.

38. Bull FC, Al-Ansari SS, Biddle S, Borodulin K, Buman MP, Cardon G, et al. World Health Organization 2020 guidelines on physical activity and sedentary behaviour. Br J Sports Med. déc 2020;54(24):1451-62.

39. Malouin F, Richards CL, Jackson PL, Lafleur MF, Durand A, Doyon J. The Kinesthetic and Visual Imagery Questionnaire (KVIQ) for assessing motor imagery in persons with physical disabilities: a reliability and construct validity study. J Neurol Phys Ther. mars 2007;31(1):20-9.

40. Peirce J, Gray JR, Simpson S, MacAskill M, Höchenberger R, Sogo H, et al. PsychoPy2: Experiments in behavior made easy. Behav Res Methods. févr 2019;51(1):195-203.

41. Peirce J, Hirst R, MacAskill M. Building experiments in PsychoPy. 2nd edition. Los Angeles London New Delhi Singapore Washington DC Melbourne: SAGE; 2022. 299 p.

42. Granot M, Weissman-Fogel I, Crispel Y, Pud D, Granovsky Y, Sprecher E, et al. Determinants of endogenous analgesia magnitude in a diffuse noxious inhibitory control (DNIC) paradigm: do conditioning stimulus painfulness, gender and personality variables matter? Pain. mai 2008;136(1-2):142-9.

43. Murphy E, King EA. Testing the accuracy of smartphones and sound level meter applications for measuring environmental noise. Applied Acoustics. 1 mai 2016;106:16-22.

44. Morin C, Bushnell MC. Temporal and qualitative properties of cold pain and heat pain: a psychophysical study. Pain. 1 janv 1998;74(1):67-73.

45. Harwell MR. Choosing Between Parametric and Nonparametric Tests. Journal of Counseling & Development. 1988;67(1):35-8.

46. Serdar CC, Cihan M, Yücel D, Serdar MA. Sample size, power and effect size revisited: simplified and practical approaches in pre-clinical, clinical and laboratory studies. Biochem Med (Zagreb). 15 févr 2021;31(1):010502.

47. Neige C, Lebon F, Mercier C, Gaveau J, Papaxanthis C, Ruffino C. Pain, No Gain: Acute Pain Interrupts Motor Imagery Processes and Affects Mental Training-Induced Plasticity. Cereb Cortex. 22 janv 2022;32(3):640-51.

48. Cuenca-Martínez F, Suso-Martí L, León-Hernández JV, La Touche R. The Role of Movement Representation Techniques in the Motor Learning Process: A Neurophysiological Hypothesis and a Narrative Review. Brain Sciences. janv 2020;10(1):27.

49. Lim RY, Ang KK, Chew E, Guan C. A Review on Motor Imagery with Transcranial Alternating Current Stimulation: Bridging Motor and Cognitive Welfare for Patient Rehabilitation. Brain Sciences. nov 2023;13(11):1584.

50. Talukdar U, Hazarika SM, Gan JQ. Motor imagery and mental fatigue: inter-relationship and EEG based estimation. J Comput Neurosci. févr 2019;46(1):55-76.

51. Jacquet T, Lepers R, Poulin-Charronnat B, Bard P, Pfister P, Pageaux B. Mental fatigue induced by prolonged motor imagery increases perception of effort and the activity of motor areas. Neuropsychologia. 8 janv 2021;150:107701.

52. Berryman C, Stanton TR, Jane Bowering K, Tabor A, McFarlane A, Lorimer Moseley G. Evidence for working memory deficits in chronic pain: A systematic review and meta-analysis. PAIN®. 1 août 2013;154(8):1181-96.

53. Hollins M, Harper D, Gallagher S, Owings EW, Lim PF, Miller V, et al. Perceived intensity and unpleasantness of cutaneous and auditory stimuli: An evaluation of the generalized hypervigilance hypothesis. PAIN®. 1 févr 2009;141(3):215-21.

54. Mirz F, Gjedde A, Sdkilde-Jrgensen H, Pedersen CB. Functional brain imaging of tinnitus-like perception induced by aversive auditory stimuli. NeuroReport. 28 févr 2000;11(3):633.

55. Ravat S, Olivier B, Gillion N, Lewis F. Laterality judgment performance between people with chronic pain and pain-free individuals. A systematic review and meta-analysis. Physiother Theory Pract. déc 2020;36(12):1279-99.

56. Coslett HB, Medina J, Goodman DK, Wang Y, Burkey A. Can they touch? A novel mental motor imagery task for the assessment of back pain. Front Pain Res [Internet]. 5 févr 2024 [cité 17 févr 2025];4. Disponible sur: https://www.frontiersin.org/journals/pain-research/articles/10.3389/fpain.2023.1189695/full

57. Beccherle M, Scandola M. How pain and body representations transform each other: A narrative review. Journal of Neuropsychology [Internet]. [cité 17 févr 2025];n/a(n/a). Disponible sur: https://onlinelibrary.wiley.com/doi/abs/10.1111/jnp.12390

58. Riquelme-Aguado V, Di-Bonaventura S, González-Álvarez ME, Zabarte-Del Campo A, Fernández-Carnero J, Gil-Crujera A, et al. How Does Conditioned Pain Modulation Influence Motor Imagery Processes in Women with Fibromyalgia Syndrome? A Cross-Sectional Study Secondary Analysis. Journal of Clinical Medicine. janv 2024;13(23):7339.

59. Scandola M, Pietroni G, Landuzzi G, Polati E, Schweiger V, Moro V. Bodily Illusions and Motor Imagery in Fibromyalgia. Front Hum Neurosci [Internet]. 20 janv 2022 [cité 17 févr 2025];15. Disponible sur: https://www.frontiersin.org/journals/human-neuroscience/articles/10.3389/fnhum.2021.798912/full

60. Özün Öİ, Öztürk M, Üzelpasacı E. Do menstrual symptoms affect motor imagery skills in young women? Arch Gynecol Obstet [Internet]. 31 janv 2025 [cité 17 févr 2025]; Disponible sur: 10.1007/s00404-025-07936-5

61. Wriessnegger SC, Bauernfeind G, Kurz EM, Raggam P, Müller-Putz GR. Imagine squeezing a cactus: Cortical activation during affective motor imagery measured by functional near-infrared spectroscopy. Brain and Cognition. 1 oct 2018;126:13-22.

62. Chang P, Arendt-Nielsen L, Chen AC. Differential cerebral responses to aversive auditory arousal versus muscle pain: specific EEG patterns are associated with human pain processing. Exp Brain Res. 1 déc 2002;147(3):387-93.

63. Yap BWD, Lim ECW. The Effects of Motor Imagery on Pain and Range of Motion in Musculoskeletal Disorders: A Systematic Review Using Meta-Analysis. Clin J Pain. janv 2019;35(1):87-99.

64. Liechti S, Tseli E, Taeymans J, Grooten W. Prognostic Factors for Quality of Life After Interdisciplinary Pain Rehabilitation in Patients with Chronic Pain—A Systematic Review. Pain Medicine. 1 janv 2023;24(1):52-70.

65. Agerbeck AH, Martiny FHJ, Jauernik CP, Bruun KD, Rahbek OJ, Bissenbakker KH, et al. Validity of Current Assessment Tools Aiming to Measure the Affective Component of Pain: A Systematic Review. PROM. 6 juill 2021;12:213-26.

66. Subirats L, Allali G, Briansoulet M, Salle JY, Perrochon A. Age and gender differences in motor imagery. Journal of the Neurological Sciences. août 2018;391:114-7.

67. Muto H, Suzuki M, Sekiyama K. Advanced aging effects on implicit motor imagery and its links to motor performance: An investigation via mental rotation of letters, hands, and feet. Front Aging Neurosci [Internet]. 17 nov 2022 [cité 26 avr 2024];14. Disponible sur: https://www.frontiersin.org/articles/10.3389/fnagi.2022.1025667

68. Saimpont A, Malouin F, Tousignant B, Jackson PL. Assessing motor imagery ability in younger and older adults by combining measures of vividness, controllability and timing of motor imagery. Brain Research. 9 févr 2015;1597:196-209.

69. Thornton C, Baird A, Sheffield D. Athletes and Experimental Pain: A Systematic Review and Meta-Analysis. J Pain. juin 2024;25(6):104450.

70. Debarnot U, Sperduti M, Di Rienzo F, Guillot A. Experts bodies, experts minds: How physical and mental training shape the brain. Frontiers in Human Neuroscience [Internet]. 2014 [cité 25 mai 2022];8. Disponible sur: https://www.frontiersin.org/article/10.3389/fnhum.2014.00280

71. Boonstra AM, de Vries SJ, Veenstra E, Tepper M, Feenstra W, Otten E. Using the Hand Laterality Judgement Task to assess motor imagery: a study of practice effects in repeated measurements. Int J Rehabil Res. sept 2012;35(3):278-80.

72. Greiner J, Schoenfeld MA, Liepert J. Assessment of mental chronometry (MC) in healthy subjects. Archives of Gerontology and Geriatrics. mars 2014;58(2):226-30.

73. Mulder Th. Motor imagery and action observation: cognitive tools for rehabilitation. J Neural Transm. oct 2007;114(10):1265-78.

74. Kim YK, Park E, Lee A, Im CH, Kim YH. Changes in network connectivity during motor imagery and execution. PLOS ONE. 11 janv 2018;13(1):e0190715.

75. Hanakawa T, Dimyan MA, Hallett M. Motor Planning, Imagery, and Execution in the Distributed Motor Network: A Time-Course Study with Functional MRI. Cerebral Cortex. déc 2008;18(12):2775-88.

76. Fields HL. How expectations influence pain. PAIN. sept 2018;159:S3.

77. Karos K, Alleva JM, Peters ML. Pain, Please: An Investigation of Sampling Bias in Pain Research. J Pain. juill 2018;19(7):787-96.

78. Sanderson A, Wang SF, Elgueta-Cancino E, Martinez-Valdes E, Sanchis-Sanchez E, Liew B, et al. The effect of experimental and clinical musculoskeletal pain on spinal and supraspinal projections to motoneurons and motor unit properties in humans: A systematic review. European Journal of Pain. 2021;25(8):1668-701.

79. Schoth DE, Nunes VD, Liossi C. Attentional bias towards pain-related information in chronic pain; a meta-analysis of visual-probe investigations. Clinical Psychology Review. 1 févr 2012;32(1):13-25.

80. Ford B, Halaki M, Diong J, Ginn KA. Acute experimentally-induced pain replicates the distribution but not the quality or behaviour of clinical appendicular musculoskeletal pain. A systematic review. Scand J Pain. 27 avr 2021;21(2):217-37.

